# Sex Differences in Neural Activity during Worry Induction and Reappraisal in Late Life

**DOI:** 10.64898/2026.09.10.26362647

**Authors:** Leigh B. Pearcy, Ana Paula Costa, Helmet T. Karim, Dana L. Tudorascu, Carmen Andreescu

## Abstract

**Background:** Severe worry – a symptom of mood and anxiety disorders – is more prevalent in women than men across the lifespan. The neural processes contributing to this difference remain unclear, especially in older adults where severe worry is related to higher cerebrovascular and neurodegenerative processes. In this study, we analyzed regional brain activation during an in-scanner worry induction task to elucidate the neural basis of worry-related sex differences among older adults.

**Methods:** 131 older adults completed a fMRI worry induction and reappraisal task. Voxel-wise t-tests were used to compare sex differences across contrasts. Activity from regions with voxel-wise differences were analyzed using regression to determine the effect of worry severity and cognitive regulation strategies.

**Results:** Greater reappraisal use was associated with less severe worry. Compared with women, men used suppression more frequently and reported lower in-scanner worry levels following worry induction. Women had greater activity than men on the worry-neutral contrast in the bilateral cerebellum declive, left middle occipital gyrus, right cuneus, and left precuneus; and on the worry-fixation contrast in the putamen, hippocampus, ventral posterolateral nucleus of the thalamus, superior temporal, amygdala, ventral lateral nucleus of the thalamus, and pallidum. Activation in the left amygdala, left precuneus, and left middle occipital were differentially associated by sex with use of reappraisal.

**Conclusions:** Differences in brain activity reveal older women recruit regions related to complex visual tasks and emotion regulation. These regions were previously associated with worry induction and maintenance networks, supporting the higher prevalence of chronic, difficult-to-control worry among older women.

## 1 Introduction

Anxiety disorders are chronic, disabling conditions affecting more than 53 million older adults worldwide, with global prevalence projected to increase an additional 20 million by 2045 (Deng et al., 2026; Grenier et al., 2019; Wang et al., 2025). Anxiety disorders contribute to an estimated 5.83 million disability-adjusted life years (DALYs) and disproportionately affect women, who experience nearly double the disability burden compared to men (Grenier et al., 2019; Wang et al., 2025). Among late-life anxiety disorders, generalized anxiety disorder (GAD) has as key symptom severe, persistent, and difficult-to-control worry. Late-life anxiety and severe worry have been associated with increased risk of coronary heart disease (Denollet et al., 2009; Emdin et al., 2016) and stroke (Lambiase et al., 2014). Recent studies reported on the association between anxiety and an increased risk of Alzheimer’s disease and related dementias (ADRD), independent of depression (Becker et al., 2018; Gulpers et al., 2016). Sex differences are well established in anxiety and severe worry, with women consistently displaying higher levels of worry compared to men. Despite the clinical significance of these sex differences and the profound health consequences associated with late-life anxiety and worry, little is known about the neural mechanisms underlying the differential expression of severe worry across sexes in older adults.

Several functional magnetic resonance imaging (fMRI) studies have investigated both activation and functional connectivity among various regions involved in GAD in older adults (Andreescu et al., 2015a, 2011; Karim et al., 2025, 2017; Mochcovitch et al., 2014). Prior work from our group introduced a naturalistic in-scanner paradigm targeting worry induction and reappraisal, revealing three distinct neural systems involved in worry induction: an imagery-based emotion generation hub, a thalamo-caudate subcortical relay related to worry maintenance, and an implicit regulatory network (vmPFC, dACC, and SMA; supplementary motor area), while reappraisal recruits a distinct prefrontal-thalamic-occipital circuit (Karim et al., 2025, 2017). Worry maintenance, reflected through a lack of habituation after induction, was instead associated with a network of regions in the limbic system including the bilateral thalamus, hippocampus, and amygdala (Karim et al., 2017). Worry severity and maintenance are core components of the pathological worry experienced in anxiety and depressive disorders, and results suggest they may be distinguished by persistent limbic activation. The same task was used in studies exploring anxiety and worry in adolescents and young adults, which showed that worry induction increased activity in the paraventricular nucleus (PVN) of the hypothalamus while worry reappraisal reduced amygdala-PVN connectivity (Westbrook et al., 2026). We have previously shown that amygdala-PVN connectivity was elevated during reappraisal among older adults with GAD (Andreescu et al., 2015b), suggesting there may be distinct age-related neural responses to worry regulation. Further, older men were shown to have reduced worry severity when using an external locus of control in reappraisal statements compared to older women (Mizuno et al., 2022). Beyond these advances, a critical gap persists: the extent to which sex moderates the neural basis of worry induction and reappraisal in older adults has yet to be investigated.

Emotional regulation strategies like suppression and reappraisal are often used to regulate worry. It has been reported that women more often use cognitive reappraisal while men engage more frequently in emotional suppression, a result which holds for older adults (Preston et al., 2022). Prior studies have investigated the affect of emotional regulation on brain activity in men and women. Young adult women were reported to have higher prefrontal cortex (PFC) activity and differential patterns of amygdala activity during emotion regulation compared to men (Min et al., 2023a). A different study investigating the neural basis of reappraisal found similar results, where young men had a smaller comparative increase in PFC activity, a greater decrease in amygdala activity, and lesser engagement with ventral striatal regions than women (McRae et al., 2008). Another study reported that, while young adult women recruited the amygdala more than men in response to negative visual stimuli, no sex-differences in amygdala activity were found during emotion down-regulation (Domes et al., 2010).

A more complete understanding of sex differences in the neural response to anxiety, stress, or worry is pivotal to develop personalized treatments. Results from previous fMRI studies point towards a differential use of neural resources between men and women when inducing emotional states such as anxiery or worry. Women were found to recruit brain regions associated with visual processing more often than men during anxiety induction, while men showed increased activity in the caudate and thalamus (Seo et al., 2017). Additionally, adolescent and young adult females had higher activity in the amygdala and PVN of the hypothalamus, results linked to the heightened somatic symptoms of anxiety in young women (Westbrook et al., 2026). Another study found that postpubertal adolescent females endorsed higher levels of trait anxiety than males mediated by higher blood perfusion in the left amygdala (Kaczkurkin et al., 2016). Sex differences in neural activation during in-scanner emotional regulation have been reported in various studies. Women showed increased activation in limbic regions and areas related to emotion processing including the right superior temporal gyrus and right middle temporal gyrus during stress induction compared to men while also reporting higher subjective stress after regulation (Kogler et al., 2015), results similar to a previous study that suggests higher stress levels, measured using self-report and cerebral blood flow, were related to greater amygdala activation in women but not men (Wang et al., 2007).

In this study, we investigated the neural basis of worry induction and worry reappraisal in older men versus older women, and examine how different anxiety phenotypes (including worry, global anxiety, rumination) and emotional regulation strategies (expressive suppression and cognitive reappraisal) modulate regional brain activation during an in-scanner worry paradigm. We hypothesized that (1) there will be significant sex differences in brain activity within regions previously found to be associated with worry maintenance (Karim et al., 2017), a result which may contribute to the increased prevalence of GAD in women, and (2) anxiety-related phenotypes would differentially predict neural activation in older men and women, reflecting potentially distinct neurobiological mechanisms in worry induction.

## 2 Methods

### 2.1 Participants and Study Design

Participants were recruited for the Functional Neuroanatomy Correlates of Worry in Older Adults (FINA) study (R01 MH108509) through in-person referrals, flyers, radio and television ads, and a web-based recruitment platform at the University of Pittsburgh called Pitt+Me. The FINA Study was approved by the University of Pittsburgh Institutional Review Board and all participants provided written informed consent prior to enrollment.

In total, we recruited 131 participants aged 50 years and older. Individuals were recruited according to their worry severity measured using the Penn State Worry Questionnaire (PSWQ) (Meyer et al., 1990). A dimensional recruitment approach allowed us to recruit participants across a wide range of worry severity. Diagnosed anxiety disorders (generalized anxiety disorder, panic disorder, etc.) and/or depressive disorders (major depressive disorder, persistent depressive disorder, etc.) were permissible, and categorical diagnoses were assessed using the structured clinical interview for DSM-V (SCID-V).

Exclusion criteria were autism spectrum disorders, intellectual development disorder, any form of psychosis or bipolar disorder, major neurocognitive disorders (e.g., dementia), and personality disorders. We excluded participants who had a modified mini-mental exam (3MSE)(Teng and Chui, 1987) < 84, high suicide risk, use of antidepressants within the last 5-14 days, history of drug or alcohol abuse within the last 6 months, were on high doses of benzodiazepines (>2mg lorazepam), had uncorrected vision problems, less than a 6th grade reading level, or had clinical diagnoses of cerebrovascular accident, multiple sclerosis, vasculitis, or significant head trauma. Additionally, participants were excluded if they were pregnant, had ferromagnetic objects in the body, claustrophobia, or were too large to fit in the MR scanner.

The primary study psychiatrist (CA) established if an antidepressant washout period was appropriate. Washout periods typically lasted two weeks (six weeks for fluoxetine). Low dose psychotropics taken for pain, sleep, or medical conditions not including mood or anxiety disorders were allowed in most cases. Three participants using psychotropics were tapered off for the MRI.

### 2.2 Assessments

We assessed worry severity using the PSWQ (Meyer et al., 1990), overall anxiety severity using the Hamilton Anxiety Rating Scale (HARS) (Hamilton, 1959), rumination severity from the Rumination Subscale from the Response Styles Questionnaire (RSQ) (Bagby et al., 2004), depression severity using the Montgomery-Asberg Depression Rating Scale (MADRS) (Montgomery and Åsberg, 1979), and habitual use of cognitive reappraisal and expressive suppression using the Emotion Regulation Questionnaire (ERQ-reappraisal, ERQ-suppression) (Gross and John, 2003). We computed the brooding and reflection components of rumination from the RSQ and additionally assessed the following domains: neuroticism (NEO Five-Factor Inventory) (Costa, Jr, 1992), perceived stress (Cohen’s Perceived Stress Scale, PSS) (Cohen, 1988), cumulative illness severity (Cumulative Illness Rating Scale for Geriatrics, CIRS-G) (Salvi et al., 2008), and cognitive function (3MSE). Participants underwent neurocognitive testing using the Repeatable Battery for the Assessment of Neuropsychological Status (RBANS)(Randolph et al., 1998) where the following domains were assessed: attention index score, language index score, immediate memory index score, delayed memory index score, and total cognitive function. We assessed executive function using the Delis-Kaplan Executive Function System (DKEFS) trail making conditions 4 vs. 5 (scaled) [set shifting]. An expert neuropsychologist with the study adjudicated participants for mild cognitive impairment (MCI).

### 2.3 Worry Induction and Reappraisal Task

We have previously published on the results from the naturalistic worry induction task used in this work to elicit in-scanner personalized worry and reappraisal (Karim et al., 2025, 2017). Prior to MRI, participants formulated personalized worry statements and rated their level of worry after reading each statement using a Likert scale from 1 (not worried at all) to 5 (extremely worried). Each participant generated at least 16 worry statements, and together with the interviewer, developed at least 8 corresponding reappraisal statements that would help regulate the worry. For example, a worry statement could be, “Worry about your husband’s health,” and a reappraisal statement could be, “Your husband has an excellent medical team.” The participant rated each worry statement to ensure they induced sufficient levels of worry (rating > 3) and each reappraisal statement to determine their effectiveness at reducing worry.

During fMRI, we presented the participants with a blocked design task where they were shown a statement for 25 seconds followed by a 10-second fixation cross. In the last 15 seconds of each statement block, participants were asked to rate their level of in-scanner worry from 1 (not worried at all) to 5 (extremely worried) using a five-finger MR-compatible glove. Statement blocks included either worry, reappraisal, or neutral statements. Neutral statements were fact-based, non-worrisome statements like, “There are 26 letters in the alphabet.” Reappraisal blocks always immediately followed their paired worry block. The 20-minute task consisted of 16 worry blocks, 8 reappraisal blocks, and 10 neutral blocks. The order of these blocks was randomized to four different scan orders, to which participants were randomly assigned. The task was implemented using Psychtoolbox v.3 in MATLAB.

### 2.4 MRI Acquisition

Scans were conducted at the MR Research Center at the University of Pittsburgh using 3T Siemens MAGNETOM Prisma scanner and a 32-channel head coil. A sagittal, whole-brain T1-weighted magnetization prepared rapid gradient echo (MPRAGE) was collected with repetition time (TR)=2400ms, echo time (TE)=2.22ms, flip angle (FA)=8deg, field of view (FOV)=320×300 with 208 slices, 0.8mm^3^ isotropic resolution, 0.4mm slice gap, and GeneRalized Autocalibrating Partial Parallel Acquisition (GRAPPA) with acceleration factor of 2 yielding a total time of 6.63 minutes. A sagittal, whole-brain T2-weighted Sampling Perfection with Application optimized Contrasts using different flip angle Evolution (SPACE) was collected with TR=3200ms, TE=563ms, FA=120deg, FOV=320×300 with 208 slices, 0.8mm^3^ isotropic resolution, no slice gap, and GRAPPA with acceleration factor of 2, with a total time of 5.95 minutes. An axial, whole-brain T2-weighted Fluid Attenuated Inversion Recovery (FLAIR) was collected with TR=10,000ms, TE=91ms, FA=135deg, inversion time (TI)=2,500ms, FOV=320×320 with 104 slices, 0.8mm×0.8mm×1.6mm resolution, no slice gap, and GRAPPA with acceleration factor of 2, with a total time of 5.95 minutes. Whole brain T2*-weighted blood oxygen level-dependent (BOLD) images were collected for a total of 20 minutes using a gradient-echo echoplanar imaging sequence in axial orientation with TR=1000ms, TE=30ms, matrix size=96×96, voxel size=2.3mm^3^ isotropic (2.3mm slice gap), FA=45 degrees, and multiband acceleration of 5. In total, participants were in the MR scanner for approximately 45-60 minutes.

### 2.5 Structural and Functional MRI Processing

We conducted structural and functional processing in SPM12 (Penny et al., 2011). Structural coregistration was performed between the MPRAGE and each of the structural images independently. We then conducted multispectral segmentation to segment to 6 tissue classes and generate a deformation field that can be used to normalize images to a standard anatomical space (Montreal Neurological Institute or MNI) (Ashburner and Friston, 2005). We created an intracranial volume mask by thresholding the gray matter, white matter, and cerebrospinal fluid tissue probability maps by 0.1 and then conducting image filling and image closing in MATLAB. We then removed the skull of the MPRAGE using this mask.

For functional images, we conducted motion correction, skull stripping (using brain extraction tool, BET), coregistration to the skull-stripped MPRAGE, normalization to MNI space, and spatial smoothing with a full-width at half-maximum (FWHM) of 8mm. We then convolved boxcars of the timings of worry, reappraisal, and neutral blocks with the canonical hemodynamic response function as input into a general linear model to identify regions with activation during those specific tasks while adjusting for the mean signal and 6 rigid body parameters of motion. We included a high-pass filter (1/128 Hz to account for drift) as well as an autoregressive [AR(1)] model to account for serial correlations due to aliased biorhythms or unmodeled activity. We specifically modeled activation for worry, reappraisal, and neutral conditions. Main effects of the task comparing worry, reappraisal, and neutral have been previously reported (Karim et al., 2025).

### 2.6 Statistical Analysis

One participant was excluded from the analyses due to missing in-scanner behavioral data (worry ratings) and 2 were excluded due to high levels of in-scanner motion (>30% of volumes a framewise displacement >0.5 mm) (Power et al., 2014). We performed voxel-wise statistical analyses in SnPM toolbox (Nichols and Holmes, 2002). We conducted independent t-tests comparing men and women on the following contrasts, adjusting for age: worry–fixation, reappraisal–fixation, worry–neutral, reappraisal–neutral, and reappraisal–worry. We used SnPM to perform permutation testing (10,000 permutations) and used a cluster-forming threshold (p = 0.001) with cluster-wise inference to control the family-wise error rate α = 0.05/5 contrasts = 0.01. We computed non-parametric p-values as these have been found to be valid for any spatial autocorrelation, while traditional p-values may lead to biased type I error control (Eklund et al., 2016). Worry–fixation and worry–neutral contrasts showed significant sex differences (see Results). We extracted brain activity from regions of interest (ROIs) with significant sex-differences on these contrasts and conducted subsequent statistical analyses.

The following linear regression analyses were implemented in R (version 4.4.1). First, regression models were used to compare sex differences in anxiety (HARS), worry (PSWQ), RSQ-brooding, RSQ-reflection, ERQ-reappraisal, and ERQ-suppression, adjusting for age. We also analyzed sex differences in worry ratings during worry, reappraisal and neutral blocks, adjusting for age. Second, we created linear regression models to explore the relationship between brain activity from ROIs with anxiety phenotypes (PSWQ, HARS, RSQ) and regulation strategies (ERQ-suppression, ERQ-reappraisal). A different model was used for each ROI, and all models adjusted for age, sex, race, education level, CIRS-G, MADRS, and neuroticism. A false discovery rate (FDR) correction for multiple comparisons was applied for each contrast separately. The residuals were examined and no assumption violations were detected.

Lastly, we performed exploratory analyses on ROI activation that included sex-based interaction terms with either PSWQ, HARS, RSQ, ERQ-suppression, or ERQ-reappraisal. A separate model was used to analyze each interaction. All models linearly adjusted for each of these anxiety phenotypes and regulation strategies, as well as other features including age, sex, race, education, CIRS-G, MADRS, and neuroticism. Partial F-tests were used to determine whether the interaction term significantly improved the model fit.

## 3 Results

### 3.1 Sample demographics

The study sample consisted of 131 participants with a mean age of 61.2 ± 8.54 years, and 61.8% of participants were women. There were no statistical differences in age, education level, or race across groups based on sex. ERQ-suppression and RSQ-reflection scores differed by sex, though the difference in RSQ-reflection did not persist after age-adjustment. Worry severity was distributed as designed, with an average PSWQ score of 51 and a range between 16 and 80. The demographic and clinical characteristics of the study sample are shown in Table 1.

**Table 1:**
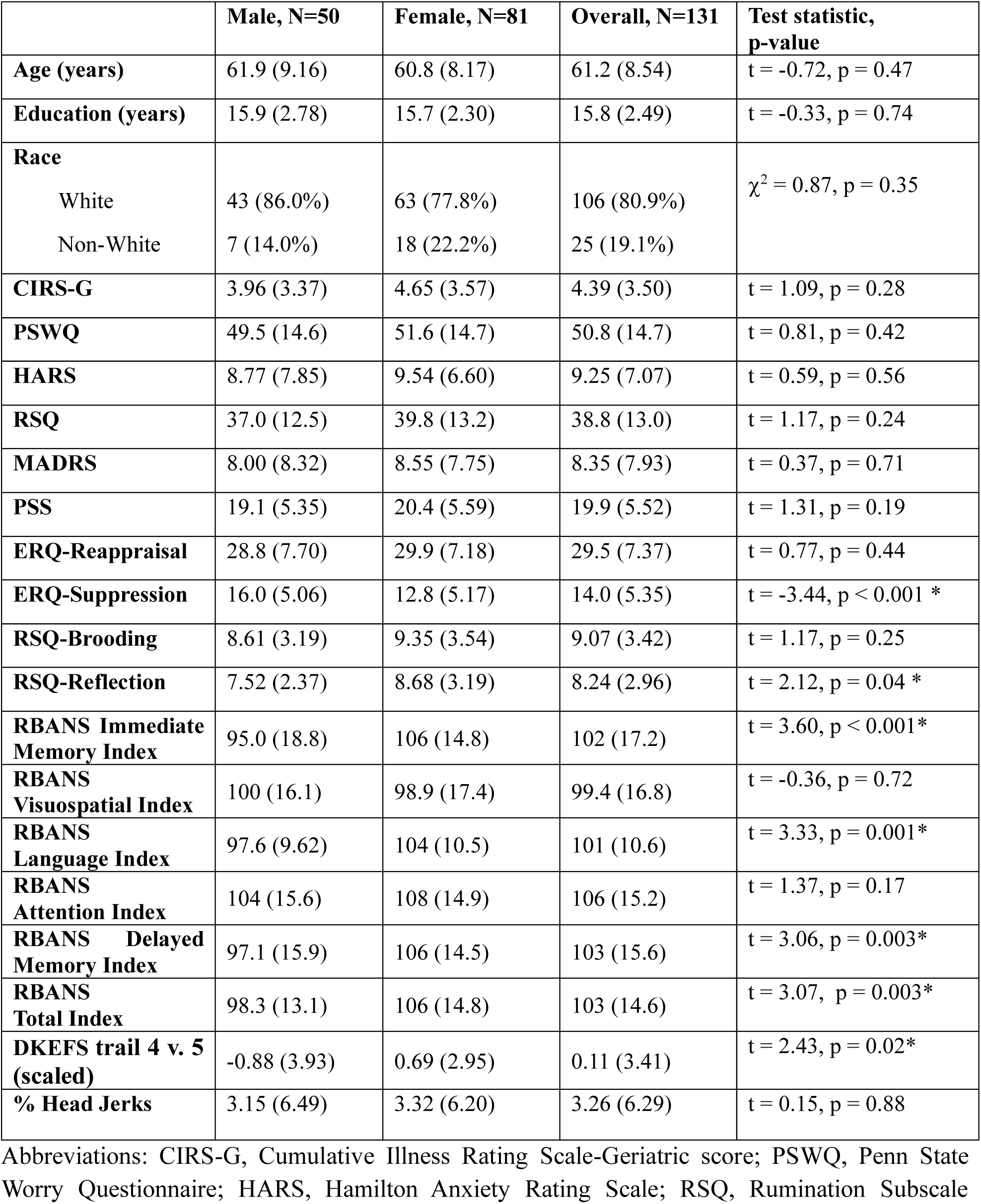
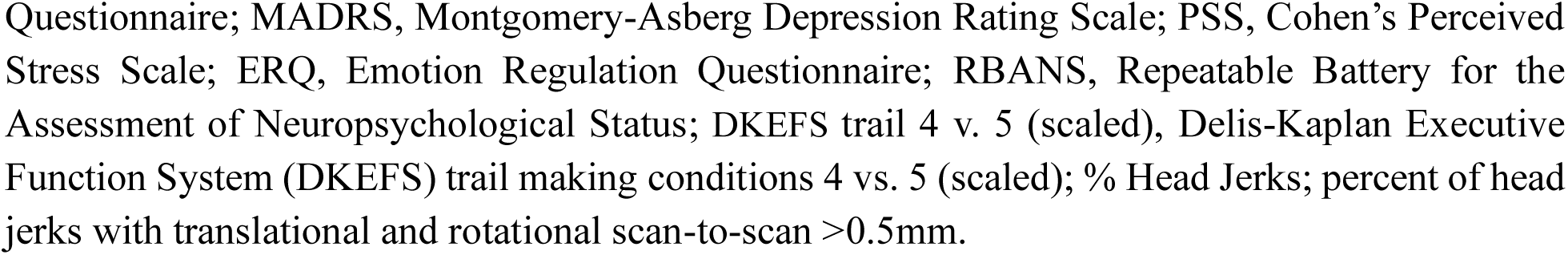
Clinical and demographic characteristics of the sample included in this analysis. Values in the table represent either the mean (standard deviation, SD) or # (%). We performed a Pearson’s Chi-squared test or a Welch’s t-test to compute test statistics and p-values.

|  | Male, N=50 | Female, N=81 | Overall, N=131 | Test statistic, p-value |
| --- | --- | --- | --- | --- |
| **Age (years)** | 61.9 (9.16) | 60.8 (8.17) | 61.2 (8.54) | t = -0.72, p = 0.47 |
| **Education (years)** | 15.9 (2.78) | 15.7 (2.30) | 15.8 (2.49) | t = -0.33, p = 0.74 |
| **Race** | | | | $\chi^2 = 0.87$, p = 0.35 |
| White | 43 (86.0%) | 63 (77.8%) | 106 (80.9%) |  |
| Non-White | 7 (14.0%) | 18 (22.2%) | 25 (19.1%) |  |
| **CIRS-G** | 3.96 (3.37) | 4.65 (3.57) | 4.39 (3.50) | t = 1.09, p = 0.28 |
| **PSWQ** | 49.5 (14.6) | 51.6 (14.7) | 50.8 (14.7) | t = 0.81, p = 0.42 |
| **HARS** | 8.77 (7.85) | 9.54 (6.60) | 9.25 (7.07) | t = 0.59, p = 0.56 |
| **RSQ** | 37.0 (12.5) | 39.8 (13.2) | 38.8 (13.0) | t = 1.17, p = 0.24 |
| **MADRS** | 8.00 (8.32) | 8.55 (7.75) | 8.35 (7.93) | t = 0.37, p = 0.71 |
| **PSS** | 19.1 (5.35) | 20.4 (5.59) | 19.9 (5.52) | t = 1.31, p = 0.19 |
| **ERQ-Reappraisal** | 28.8 (7.70) | 29.9 (7.18) | 29.5 (7.37) | t = 0.77, p = 0.44 |
| **ERQ-Suppression** | 16.0 (5.06) | 12.8 (5.17) | 14.0 (5.35) | t = -3.44, p < 0.001 * |
| **RSQ-Brooding** | 8.61 (3.19) | 9.35 (3.54) | 9.07 (3.42) | t = 1.17, p = 0.25 |
| **RSQ-Reflection** | 7.52 (2.37) | 8.68 (3.19) | 8.24 (2.96) | t = 2.12, p = 0.04 * |
| **RBANS Immediate Memory Index** | 95.0 (18.8) | 106 (14.8) | 102 (17.2) | t = 3.60, p < 0.001* |
| **RBANS Visuospatial Index** | 100 (16.1) | 98.9 (17.4) | 99.4 (16.8) | t = -0.36, p = 0.72 |
| **RBANS Language Index** | 97.6 (9.62) | 104 (10.5) | 101 (10.6) | t = 3.33, p = 0.001* |
| **RBANS Attention Index** | 104 (15.6) | 108 (14.9) | 106 (15.2) | t = 1.37, p = 0.17 |
| **RBANS Delayed Memory Index** | 97.1 (15.9) | 106 (14.5) | 103 (15.6) | t = 3.06, p = 0.003* |
| **RBANS Total Index** | 98.3 (13.1) | 106 (14.8) | 103 (14.6) | t = 3.07, p = 0.003* |
| **DKEFS trail 4 v. 5 (scaled)** | -0.88 (3.93) | 0.69 (2.95) | 0.11 (3.41) | t = 2.43, p = 0.02* |
| **% Head Jerks** | 3.15 (6.49) | 3.32 (6.20) | 3.26 (6.29) | t = 0.15, p = 0.88 |

Abbreviations: CIRS-G, Cumulative Illness Rating Scale-Geriatric score; PSWQ, Penn State Worry Questionnaire; HARS, Hamilton Anxiety Rating Scale; RSQ, Rumination Subscale
Questionnaire; MADRS, Montgomery-Asberg Depression Rating Scale; PSS, Cohen's Perceived Stress Scale; ERQ, Emotion Regulation Questionnaire; RBANS, Repeatable Battery for the Assessment of Neuropsychological Status; DKEFS trail 4 v. 5 (scaled), Delis-Kaplan Executive Function System (DKEFS) trail making conditions 4 vs. 5 (scaled); % Head Jerks; percent of head jerks with translational and rotational scan-to-scan >0.5mm.

### 3.2 Sex-based differences in clinical characteristics

Men had higher levels of ERQ-suppression than women and lower mean in-scanner worry levels during the worry induction blocks (Supplemental Fig. 1A). No other clinical characteristic examined (worry severity, anxiety, RSQ-brooding, RSQ-reflection, ERQ-reappraisal) differed by sex when adjusting for age. Sex differences in ERQ-suppression and mean in-scanner worry were examined separately for low worriers (PSWQ ≤ 45) and high worriers (PSWQ > 45), finding each effect was driven primarily by low worriers (Supplemental Fig. 1B). Among low worriers, men reported lower mean in-scanner worry than women and women reported lower levels of ERQ-suppresssion than men. There were no differences found in either measure among high worriers (Supplemental Fig. 1C).

No sex differences were found in mean in-scanner worry levels during the neutral and reappraisal blocks after adjusting for age (neutral: t(125)=1.5, p=0.13; reappraisal: t(125)=1.6, p=0.11). Further, there were no differences in the ability of men and women to reappraise after worry, which was measured by the change in in-scanner worry severity ratings from worry to reappraisal blocks (t(125)=0.41, p=0.68). Lastly, greater use of reappraisal (greater ERQ-reappraisal) was associated with decreased worry measured with the PSWQ (t(118)= −3.72, p < 0.001) adjusting for age and sex, while greater use of suppression (ERQ-suppression) was not (t(118)=0.55, p=0.58). See Supplemental Fig. 2.

### 3.3 Activation across brain regions

We found that women had greater activity compared to men on the worry–neutral contrast (Fig. 1, top) in the left middle occipital gyrus (t-max=3.9, x=-36, y=-72, z=24, 825 voxels), bilateral cerebellum declive (t-max=4.4, x=-4, y=-62, z=-18, 703 voxels), right cuneus (t-max=4.9, x=10, y=-100, z=18, 361 voxels), and left precuneus (t-max=4.3, x=-19, y=-53, z=43, 321 voxels). We found no sex differences on the reappraisal–neutral or reappraisal–worry contrasts.

**Figure 1.**
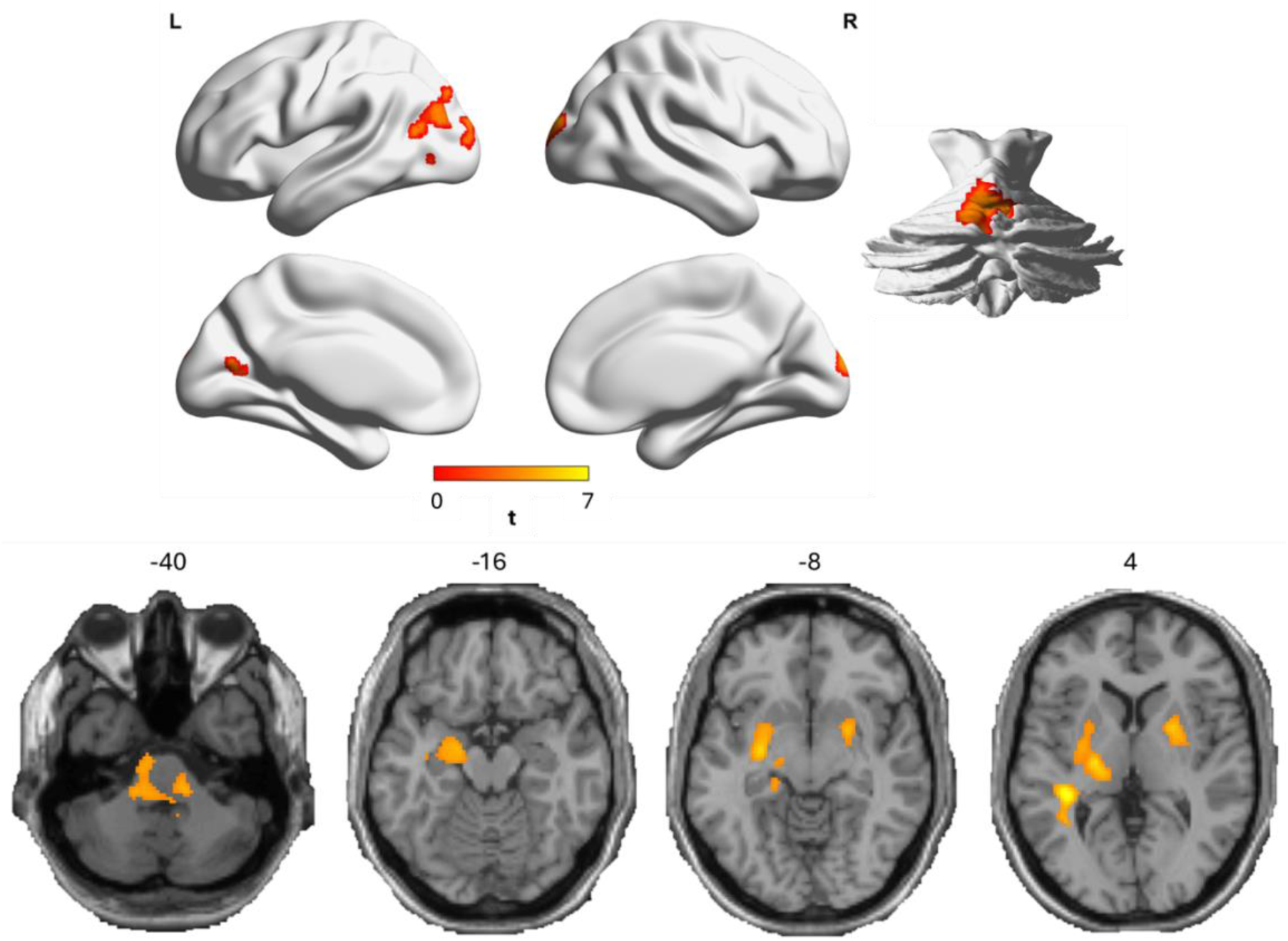
Sex differences (women > men) on Worry–Neutral contrast (top) and Worry–fixation (bottom).

We additionally found that women had greater activity compared to men on the worry–fixation contrast (Fig. 1, bottom) in the left putamen (t-max = 4.3, x = −28, y = −8, z = −2, 345 voxels), hippocampus (t-max = 4.3, x = −26, y = −10, z = −12, 147 voxels), ventral posterolateral nucleus of the thalamus (VPL; t-max = 4.2, x = −20, y = −18, z = 2, 120 voxels), pallidum (t-max = 4.1, x = −26, y = −10, z = −2, 76 voxels), superior temporal gyrus (t-max = 4.6, x = −40, y = −32, z = 4, 74 voxels), amygdala (t-max = 4.3, x = −26, y = −8, z = −12, 64 voxels), and ventral lateral nucleus of the thalamus (VL; t-max = 4.0, x = −18, y = −16, z = 2, 50 voxels), as well as the right putamen (t-max = 3.9, x = 24, y = 4, z = 6, 212 voxels) and pallidum (t-max = 3.8, x = 24, y = 2, z = −6, 59 voxels). No sex differences were found when examining the reappraisal–fixation contrast.

### 3.4 Associations between ROI activation and anxiety phenotypes

Brain activation from ROIs with significant sex-differences on the worry–fixation and worry–neutral contrasts were examined separately using linear models to determine if associations with worry, anxiety, rumination, and regulation strategies including ERQ-suppression and ERQ-reappraisal exist. We adjusted for age, sex, race, education level, CIRS-G, MADRS, and neuroticism. In total, 9 models were used to examine the worry–fixation contrast and 4 models for the worry–neutral contrast (one for each ROI). We conducted FDR correction to adjust for multiple comparisons on each contrast. We reported the standardized β coefficients in Tables 2 and 3, where significant results are bolded. Standardized β coefficents represent the amount of change in outcome variable per standard deviation change in input variable.

**Table 2:** Standardized regression coefficients (β) from linear regression models predicting worry–fixation activation across specified regions. Models controlled for age, sex, race, education, CIRS-G, MADRS, and neuroticism. Bolded β values indicate the variable (row) is significant at p = 0.05 (Wald tests, 12 df) for activation in that region (column).

|  | Left<br>Hippocampus | Left<br>Pallidum | Right<br>Pallidum | Left<br>Putamen | Right<br>Putamen | Left<br>Superior<br>Temporal<br>Gyrus | Left<br>VPL | Left<br>Amygdala | Left<br>VL |
| --- | --- | --- | --- | --- | --- | --- | --- | --- | --- |
| Age | 0.035 | -0.040 | 0.014 | 0.008 | 0.028 | 0.044 | 0.037 | -0.011 | 0.042 |
| Sex | **0.376** | **0.449** | **0.456** | **0.456** | **0.454** | **0.447** | **0.484** | **0.318** | **0.467** |
| Race | -0.151 | 0.050 | 0.042 | 0.044 | 0.019 | 0.109 | -0.051 | -0.158 | -0.076 |
| Edu | 0.035 | 0.070 | 0.117 | 0.057 | 0.097 | 0.024 | -0.030 | -0.015 | -0.017 |
| CIRS-G | 0.073 | 0.190 | 0.119 | **0.204** | 0.141 | -0.019 | 0.178 | 0.149 | 0.175 |
| MADRS | 0.230 | 0.025 | -0.097 | 0.034 | -0.059 | 0.042 | 0.051 | 0.205 | 0.000 |
| Neuroticism | -0.117 | 0.045 | 0.099 | 0.054 | 0.045 | -0.077 | 0.027 | -0.269 | 0.033 |
| RSQ | 0.158 | -0.053 | 0.067 | -0.009 | 0.099 | -0.079 | -0.026 | 0.244 | 0.002 |
| HARS | 0.009 | 0.147 | 0.269 | 0.158 | 0.230 | 0.103 | 0.018 | 0.124 | 0.107 |
| ERQ-reappraisal | -0.141 | -0.142 | -0.081 | -0.123 | -0.127 | -0.053 | -0.117 | -0.178 | -0.108 |
| ERQ-suppression | -0.089 | 0.161 | **0.221** | 0.129 | 0.168 | 0.157 | **0.258** | -0.147 | **0.270** |
| PSWQ | -0.203 | -0.264 | **-0.333** | **-0.302** | **-0.317** | 0.116 | -0.225 | -0.239 | -0.196 |

Abbreviations: df, degrees of freedom; CIRS-G, Cumulative Illness Rating Scale-Geriatric score; MADRS, Montgomery-Asberg Depression Rating Scale; RSQ, Rumination Subscale Questionnaire; HARS, Hamilton Anxiety Rating Scale; ERQ, Emotion Regulation Questionnaire; PSWQ, Penn State Worry Questionnaire.

**Table 3:** Standardized regression coefficients (β) from linear regression models predicting worry–neutral activation across specified regions. Models controlled for age, sex, race, education, CIRS-G, MADRS, and neuroticism. Bolded β values indicate the variable (row) is significant at p = 0.05 (Wald tests, 12 df) for activation in that region (column).

|  | Left Precuneus | Left Mid Occipital Gyrus | Cerebellum Declive B | Right Cuneus |
| --- | --- | --- | --- | --- |
| Age | 0.015 | 0.009 | -0.106 | -0.140 |
| Sex | **0.364** | **0.367** | **0.380** | **0.337** |
| Race | 0.096 | 0.161 | -0.181 | -0.001 |
| Edu | 0.111 | 0.016 | 0.083 | -0.105 |
| CIRS-G | -0.198 | **-0.193** | -0.189 | -0.090 |
| MADRS | 0.202 | 0.238 | 0.165 | 0.248 |
| Neuroticism | 0.125 | 0.027 | 0.134 | -0.063 |
| RSQ | -0.206 | -0.197 | -0.249 | -0.124 |
| HARS | -0.081 | 0.036 | 0.093 | -0.079 |
| ERQ-reappraisal | 0.086 | 0.145 | 0.009 | 0.049 |
| ERQ-suppression | -0.043 | -0.079 | 0.034 | **-0.205** |
| PSWQ | -0.036 | -0.062 | -0.086 | -0.001 |

Abbreviations: df, degrees of freedom; CIRS-G, Cumulative Illness Rating Scale-Geriatric score; MADRS, Montgomery-Asberg Depression Rating Scale; RSQ, Rumination Subscale Questionnaire; HARS, Hamilton Anxiety Rating Scale; ERQ, Emotion Regulation Questionnaire; PSWQ, Penn State Worry Questionnaire.

Greater worry–neutral activation in the right cuneus was associated with lower ERQ-suppression (lower habitual use of expressive suppression). Greater worry activation in the right pallidum, VPL, and VL were all associated with greater ERQ-suppression, while greater worry activation in the right pallidum, and the left and right putamen were associated with lower worry severity (PSWQ). These results are shown in Fig. 2.

**Figure 2.**
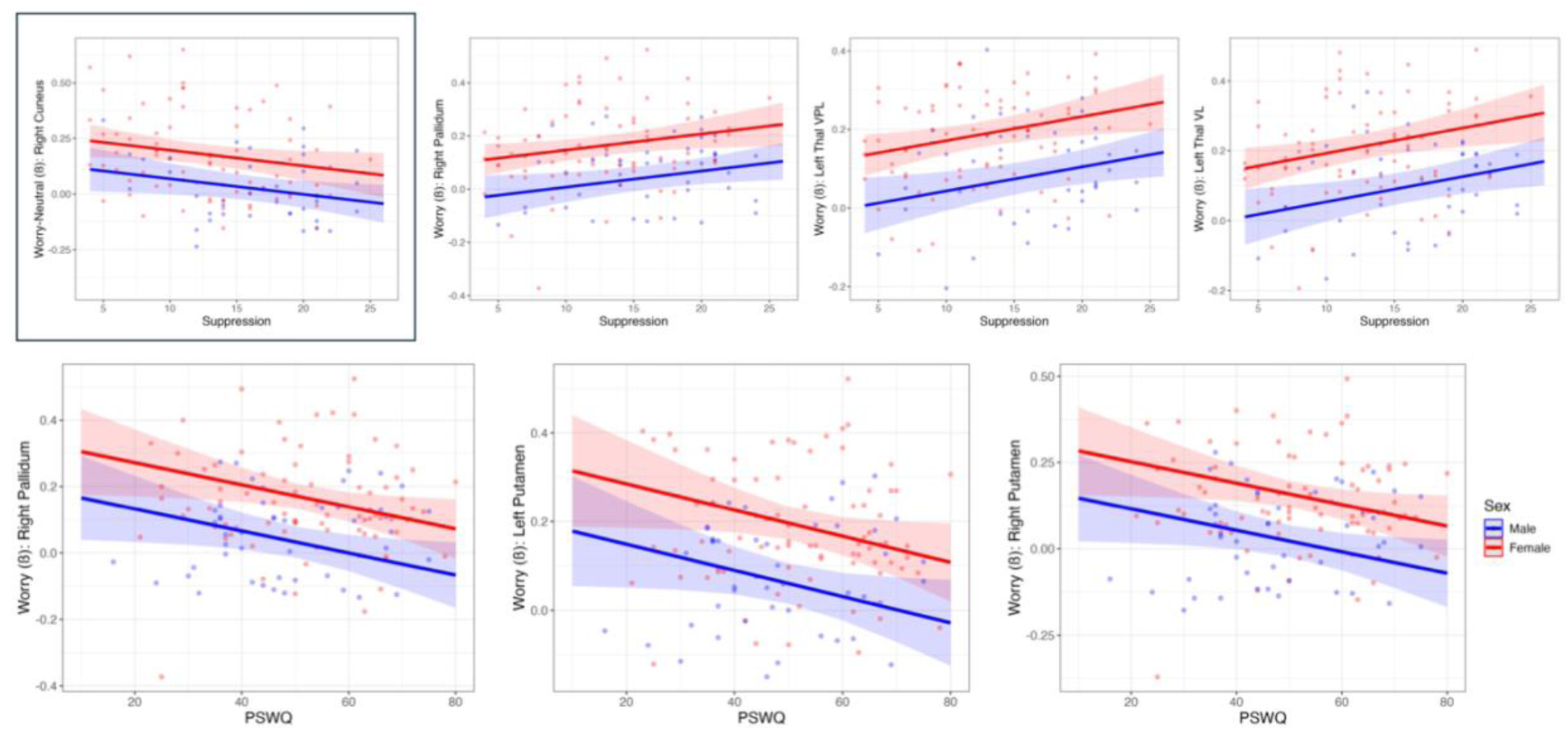
Significant effects of ERQ-suppression (top row) and PSWQ (bottom row) on activation in ROIs. The upper left plot (boxed) represents the worry–neutral contrast. All other figures result from the worry–fixation contrast.

In the exploratory analysis, interactions between sex and habitual use of reappraisal (ERQ-reappraisal) were found to improve model fit for ROI activation on both the worry–fixation and worry–neutral contrasts (Fig. 3). Specifically, greater ERQ-reappraisal (greater habitual use of reappraisal) was associated with a lower activation in the left amygdala for men on the worry–fixation contrast. We also found that greater ERQ-reappraisal was associated with greater worry–neutral activation in the left precuneus and the left middle occipital for women.

**Figure 3.**
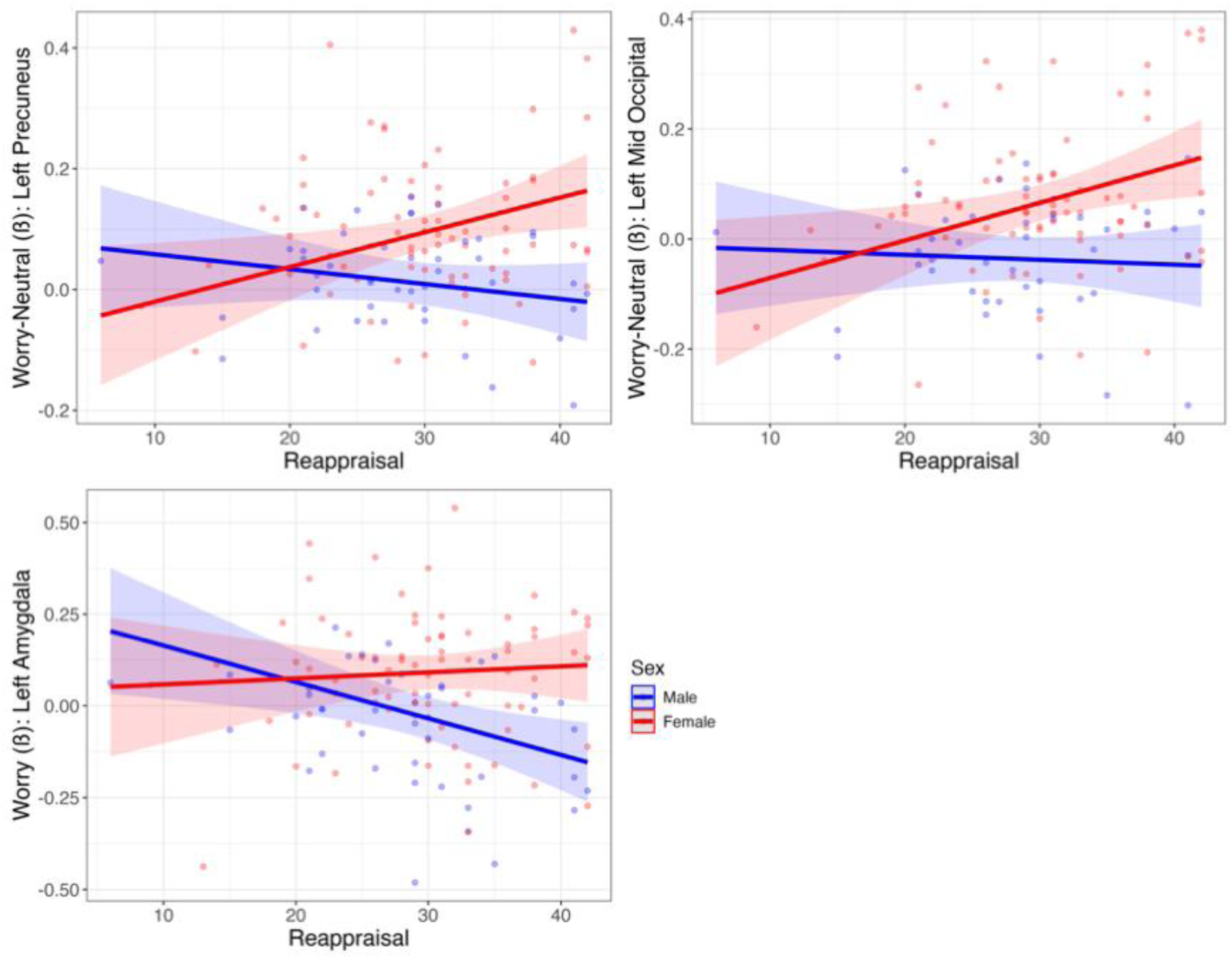
Results of the exploratory analysis examining differences in ROI activation in models including sex-based interaction terms. Greater worry–neutral activation in the left precuneus and the left middle occipital gyrus was associated with greater ERQ-reappraisal in women (top row). Greater worry–fixation activation in the left amygdala was associated with lower ERQ-reappraisal in men.

## 4. Discussion

The present study examined sex differences in the neural basis of late-life worry induction and worry reappraisal in adults 50 years and older. Men and women had similar levels of worry, anxiety, and rumination. Men were more likely to use expressive suppression. Consistent with our hypothesis, women exhibited greater brain activation than men during worry induction, particularly in regions associated with visual imagery and emotional processing - including the middle occipital cortex, precuneus, bilateral cerebellum declive, and putamen - as well as subcortical structures critically involved in emotional processing, such as the amygdala, hippocampus, thalamus, and pallidum. Importantly, no sex differences were observed for the reappraisal task contrasts, suggesting that sex-related neural differences are specific to the processes underlying worry generation rather than to the cognitive reappraisal of worry. Activation in several subcortical regions were associated with increased habitual use of expressive suppression and lower worry severity as measured by PSWQ. Exploratory analyses revealed sex-moderated associations with habitual use of cognitive reappraisal: greater ERQ-reappraisal was associated with lower left amygdala activation (worry–fixation) in men and with greater left visual cortex activation (worry–neutral) in women. Collectively, these findings extend prior work from our group(Karim et al., 2025) by demonstrating that sex is an important biological variable influencing the neural basis of worry in older adults, and underscore the importance of incorporating sex into neuroimaging models of late-life anxiety.

These findings can be interpreted within the framework of the transdiagnostic processes of repetitive negative thinking (RNT), which characterizes worry and rumination as repetitive and difficult-to-control negative thinking processes involved in the onset and maintenance of multiple emotional disorders (Clancy et al., 2016; Gustavson et al., 2018; Wahl et al., 2019). Within this framework, a useful distinction can be drawn between the initiation and maintenance of the worry processes, which aligns with the continuum worry spectrum from adaptative, mild worries to severe, difficult-to-control worry encountered in disorders such as GAD (Karim et al., 2017). Thus, more severe worry requires both a heightened activation during initial induction and a maintenance of the worry processes, with limited reduction following explicit emotion regulation strategies.

During worry induction, the greater activation observed in women within occipital and parietal regions - including the occipital cortex and precuneus - may reflect greater recruitment of neural systems involved in visual imagery, episodic retrieval, and self-referential processing. The precuneus has been consistently implicated in visuospatial imagery, episodic memory, and internally directed cognition (Cavanna and Trimble, 2006; Tanguay et al., 2023), while occipital and high-level visual cortical activation may reflect recruitment of visual systems involved in mental imagery processes - with recent evidence pointing specifically to fronto-parietal networks and the left fusiform gyrus as core substrates of visual mental imagery generation (Spagna et al., 2021). This interpretation is consistent with findings from out group, indicating that self-referential worries (e.g. worry about oneself) were associated with more severe scores on the PSWQ (Mizuno et al., 2022). Also, in the largest fMRI study of worry induction in older adults to date, Karim et al. (2025) reported that worry induction robustly activated regions spanning the default mode network, executive control network, anterior salience network, visual cortex, and subcortical structures including the basal ganglia and thalamus.

Beyond initiation, women in our sample also showed greater recruitment of subcortical structures - including the amygdala, hippocampus, putamen, pallidum, and thalamus - that are critically involved in the maintenance of the worry process through threat appraisal, affective memory encoding, and subcortical relay circuitry involved also in obsessional ideation (Li et al., 2026; Ting and Feng, 2011). These regions form part of the distributed network supporting sustained emotional reactivity and are consistently implicated in anxiety-related neural processes (Akiki et al., 2025; Gong, 2025). The results of our regression analyses support previous literature indicating that women have greater activation across various limbic structures compared to men during emotional processing (Jenkins et al., 2018; Stevens and Hamann, 2012). In our study, activation in the right pallidum was indicative of a greater habitual use of expressive suppression and reduced worry measured using the PSWQ. Taken together, these patterns suggest that initiation of worry triggers a more robust visual and subcortical circuitry in women, and the sustained subcortical engagement may reflect the “persistent and difficult-to-control” features that define the transition from normative to pathological worry. This distinction carries clinical relevance: while transient worry is adaptive, persistent and uncontrollable worry is one of the hallmarks of pathological anxiety. In older adults specifically, the population prevalence of moderate-to-severe GAD is significantly higher in women than men, a disparity that systematic reviews attribute to the convergence of biological factors including sex hormone trajectories during the menopause transition, HPA axis dysregulation, and differential amygdalo-limbic reactivity across the female lifespan (Farhane-Medina et al., 2022; Marano et al., 2026). The present neuroimaging findings may help elucidate the biological underpinnings of this well-documented sex disparity.

Exploratory analyses examining the moderating role of cognitive reappraisal revealed sex-specific associations between this emotion regulation strategy and worry-related neural activation. In men, greater habitual use of reappraisal was associated with lower amygdala activation during worry induction, suggesting that reappraisal may function as an effective top-down regulatory mechanism to reduce emotional reactivity. Of note, these findings derive from an exploratory moderation analysis - examining the interaction between sex and ERQ-reappraisal scores on worry–fixation amygdala activation - and should be interpreted with appropriate caution. Critically, no sex differences were observed in any task-based reappraisal contrasts, i.e., reappraisal–neutral, reappraisal–worry, and reappraisal–fixation contrasts, indicating that men and women did not differ in amygdala or prefrontal activation during the in-scanner reappraisal of worry condition itself. Nevertheless, the observed moderation pattern is consistent with prior fMRI literature reporting that – when using reappraisal - men show greater decreases in amygdala activation and smaller increases prefrontal cortex activation (McRae et al., 2008), suggesting a more efficacious neural engagement in men during cognitive reappraisal (McRae et al., 2008; Min et al., 2023b).

In the context of worry, our exploratory findings may have potential therapeutic implications. Cognitive restructuring and reappraisal-based techniques are core components of Cognitive Behavioral Therapy (CBT) for late-life anxiety. The most current Cochrane review of psychological treatments for anxiety in older adults concluded that CBT is more effective than minimal management in reducing anxiety and worry symptoms post-treatment, though the evidence is less certain for longer-term outcomes, and important questions about subgroup moderators - including sex - remain unaddressed (Hendriks et al., 2024). If, as our exploratory data suggest, the relationship between reappraisal and amygdala reactivity during worry differs by sex, this could contribute to variability in CBT response, though this hypothesis requires direct prospective testing before clinical conclusions can be drawn. A second, complementary finding from the exploratory moderation analyses further enriches the neural picture of women’s late-life worry. Greater habitual use of reappraisal was associated with greater worry–neutral activation in the left precuneus and left middle occipital gyrus specifically in women. This finding does not indicate that these regions were activated during the in-scanner reappraisal task condition, but rather that individual differences in habitual use of reappraisal moderated the degree of worry-related activation in visual regions in a sex-specific manner. Together with the primary finding of greater occipital and precuneus activation in women during worry induction (worry–neutral contrast), this pattern raises the possibility that, in women, severe worry and its relationship to reappraisal may be more strongly coupled to visual and imagery-based neural systems than in men. This interpretation is consistent with the broader literature on worry and mental imagery. Although the dominant characterization of worry as primarily a verbal-linguistic process(Borkovec and Inz, 1990) has shaped cognitive models of GAD, a growing body of evidence demonstrates that mental imagery plays a clinically meaningful role in worry (Dance et al., 2025; Holmes and Mathews, 2010; Stavropoulos et al., 2024). Future studies should directly test whether sex moderates response to imagery-based versus standard verbal-cognitive CBT protocols for late-life anxiety.

Our study has several limitations that should be considered when interpreting the findings. Although our sample of older adults with a broad range of worry severity represents a meaningful contribution to the neuroimaging literature on late-life anxiety and worry, the relatively modest sample size, particularly when stratified by sex for the exploratory moderation analyses, may have limited statistical power to detect smaller effects. These results will need further replication in larger samples. Another limitation is the cross-sectional design of the study precludes causal inference regarding the relationships between sex, emotion regulation, and neural activation during worry. We cannot determine whether the observed sex differences in neural activation represent stable neurobiological traits, state-dependent responses to the experimental paradigm, or the cumulative effects of lifetime differences in worry experience and emotion regulation habits. The generalizability of our findings may be limited by the demographic characteristics of the sample, which was predominantly non-Hispanic White and recruited from a single study site. Finally, although we adjusted for several relevant covariates in all analyses - including age, race, education, medical burden, depression, and neuroticism - residual confounding cannot be entirely excluded.

To summarize, the present study provides new insights into sex differences in the neural basis of worry induction and worry reappraisal in older adults. Our findings highlight sex as a critical biological variable in late-life anxiety and underscore the importance of incorporating sex-stratified analyses in future neuroimaging studies. From a clinical perspective, the present findings raise the hypothesis that sex-informed interventions targeting emotion regulation, and particularly approaches that incorporate visual imagery-based techniques, may offer therapeutic advantages for women with late-life anxiety. Future longitudinal studies with larger, more diverse samples are needed to replicate these results and to determine whether sex-specific neural profiles of worry predict differential response to psychological treatment.

## Data Availability

Data is available upon request from the principal investigators.

## Authorship statement

Conceptualization: LBP, HTK, CA; Funding Acquisition: HTK, DLT, CA; Data Analysis: LBP, HTK; Writing original draft: LBP, APC, HTK, CA; Writing reviewing and editing: LBP, APC, HTK, DLT, CA.

## Acknowledgements

We would like to thank participants for their participation in the study, the study staff from the ARGO neuroscience of aging research group, and Dr. Meryl Butters for serving as the expert neuropsychologist in the study.

## Consent Statement

All participants gave written informed consent prior to starting study procedures. This study was approved by the IRB at the University of Pittsburgh.

## Previous Presentations

This work was presented at Pitt Psychiatry Annual Research Day 2026, Pittsburgh, PA, June 11, 2026.

## Data Sharing Statement

Data is available upon request from the principal investigators.

