## Supplemental figures for "Sex Differences in Neural Activity during Worry Induction and Reappraisal in Late Life"

**Title**

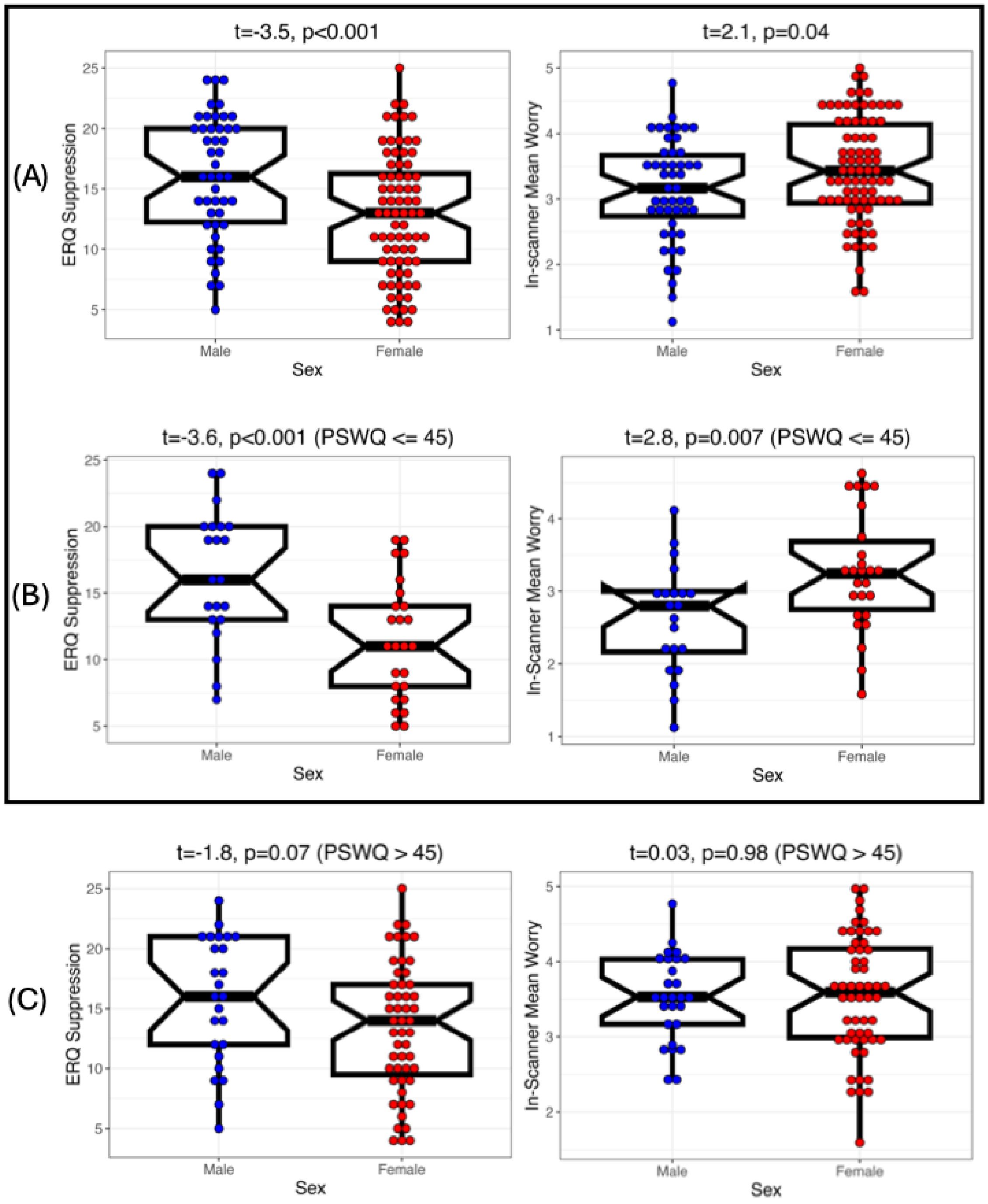


Supplemental Figure 1. (A) Sex differences in ERQ suppression (left) and in-scanner mean worry rating from the worry blocks (right) from the total sample of N = 131 participants. Sex differences among (B) low worriers, N = 49, (C) high worriers, N = 82, in ERQ suppression and in-scanner mean worry. Significant results are boxed.

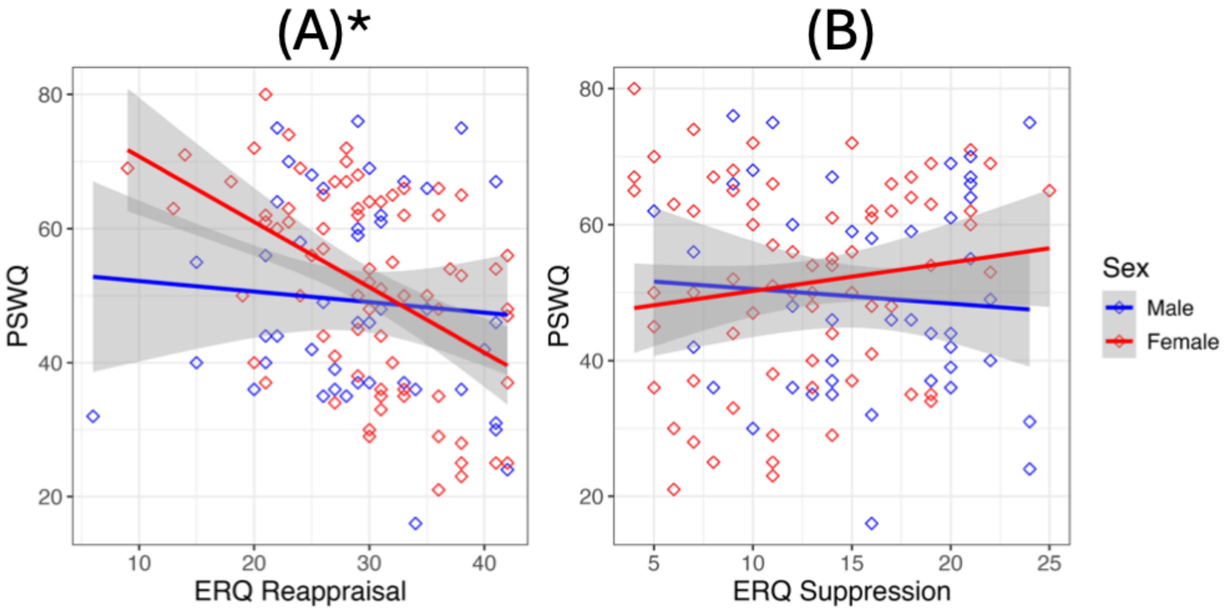


Supplemental Figure 2. (A) Relationship between worry (PSWQ) and habitual use of reappraisal (ERQ-reappraisal); star indicates significant result. Using reappraisal more often (higher ERQ-reappraisal) is associated with decreased worry (lower PSWQ) in women. (B) Relationship between worry and habitual use of suppression (not significant).
